# Clinical and Demographic Characteristics of Treatment Requiring Retinopathy of Prematurity (ROP) in Big Premature Infants in Turkey – Report No: 1 (BIG-ROP STUDY)

**DOI:** 10.1101/2022.03.14.22272395

**Authors:** Şengül Özdek, Huseyin Baran Ozdemir, Zuhal Ozen Tunay, Sadik Etka Bayramoglu, Emine A. Sukgen, Nur Kır, Esin Koç, BIGROP Study Group

## Abstract

**Aim:** To demonstrate the clinical and demographic features of infants with gestational age (GA) of 32-37 weeks (wk) and birth weight (BW) of >1500 g who developed treatment requiring retinopathy of prematurity (ROP).

**Methods:** Retrospective, observational, descriptive, multicentre study was conducted by the Turkish Ophthalmological Association ROP commission. Data on the infants with a GA of 32-37 wk and BW >1500 g who developed treatment-requiring ROP were collected from the 33 ROP centres in Turkey. GA, BW, type of hospital, neonatal intensive care units (NICU) level, length of stay in NICU, duration of oxygen therapy, comorbidities, type of ROP and time for treatment-requiring ROP (TR-ROP) development were analysed.

**Results:** Totally 366 infants were included in the study. The mean GA and BW were 33±1 wk and 1896 ± 316 g, respectively. Duration of hospitalization was 3-4 wk in 46.8% of them. The first ROP examination was performed at postnatal 4-5 wk in 80.3% of infants, which was significantly later in lower levels of NICU and non-university clinics. ROP was detected in 90.9% of infants at the first ROP examination, especially in clinics without an ophthalmologist. In 15.3% of the infants, treatment was required in postnatal fourth week, and the mean development of TR-ROP was 6.16 ± 2.04 wk

**Conclusion:** Routine ROP screening thresholds need to be expanded in hospitals with suboptimal NICU conditions considering the development of TR-ROP in more mature and heavier preterm infants, and the first ROP examination should be no later than postnatal fourth week.

**What is already known on this topic:** Treatment-requiring retinopathy of prematurity (TR-ROP) may develop in bigger and more mature infants with a gestational age >32 weeks and birth weight >1500g especially in low/middle-income countries where proper neonatal intensive care conditions could not be provided,

**What this study adds:** This is the first study analysing the regional differences and the effect of presence of the ophthalmologist and neonatologist in the same hospital as NICU on the development of TR-ROP in a nation-wide study in Turkey. This study emphasizes the high rate of ROP at the first examination in these bigger babies and progression to TR-ROP in a short time. The ROP in these infants may be more aggressive like A-ROP and may progress rapidly in a short time. The results of our study suggest the need for a timely (even earlier) screening for bigger infants and a revision to expand the limits of the ROP screening program to bigger infants in at least underdeveloped parts of Turkey. This may be generalized to all of the underdeveloped countries.

**How this study might affect research, practice or policy:** Screening criteria for ROP need to be revised for the coverage of bigger infants in Turkey depending on the NICU conditions of the hospitals. Infants with a gestational age of >32 weeks and a birth weight of >1500 g may need to be screened for ROP earlier than postnatal four weeks. Increasing the number of well-educated neonatologists and ophthalmologists as well as other NICU conditions will improve neonatal care for ROP to the standards of developed countries.

## Introduction

Retinopathy of prematurity (ROP) is a sight-threatening, vasoproliferative disease affecting peripheral immature retina in premature infants. ROP-related blindness can be potentially avoidable with advanced neonatal care and effective screening/treatment programs. The prevalence of ROP varies among countries and is influenced by the income of countries and the development of neonatal intensive care units (NICU).^1^ Treatment-requiring ROP (TR-ROP) typically occurs in larger and more mature infants in low/middle-income countries.^1^ Therefore, national screening criteria are usually set according to the demographic features of infants developing ROP.

According to Turkey ROP Screening Guideline which was established in 2016, preterm infants with gestational age (GA) ≤ 32 weeks (wk) or birth weight (BW) ≤1500 g or infants with a BW >1500 g and GA >32 wk with a defined extra-risk factor for ROP by paediatrician are screened for ROP.^2^ A previous multicentre prospective study in Turkey showed that the rates of severe ROP were higher in non-university NICUs, especially in private hospitals.^2^ The findings of the study also revealed that bigger and more mature babies are at high risk of developing severe ROP in Turkey and indicated that screening criteria should be wide enough to detect the majority of infants developing TR-ROP.^2^

In this study, we aimed to evaluate retrospectively the demographic and clinical features of TR-ROP seen in bigger infants with a GA>32 wk and BW>1500g in Turkey.

## Material and Methods

This non-randomized, retrospective, multicentre study was conducted by the Turkish Ophthalmological Association (TOA) ROP commission to evaluate the demographic and clinical features of preterm infants with a GA>32 wk and BW>1500 g who developed TR-ROP. Members of the TOA who wish to contribute were requested to fill out the study-specific data entry forms for each patient treated for TR-ROP. Submission of the fulfilled forms was made until December 2020.The study organizers (SO, HBO) received data of 713 eyes of 376 patients treated by 57 ophthalmologists from 33 centres. The institutional review board of each centre and the Ethics Commission of the Gazi University (No: 2020-191) approved the study protocol, which adhered to the tenets of the Declaration of Helsinki.

Infants with GA<32 wk or BW<1500 g, or bigger infants developing ROP who did not meet treatment criteria were excluded. Demographic data including GA, BW, city and region where the infant was born, city and region where the infant was hospitalized, level of NICU^3^, and type of hospital defined as a university hospital, state hospital, or private hospital were collected for each case.

According to the socio-economic development index map determined by the Ministry of Industry and Technology of Turkey, the cities were classified into six levels according to their development. The first three stages were evaluated as developed cities and the last three stages as underdeveloped cities in the study.^4^ Presence of neonatologist and ophthalmologist in the hospital was questioned. If there was no ophthalmologist, the details of how the ROP examination was performed were requested such as an ophthalmologist who was invited from another centre or transferring the infant to another centre only for ROP screening.

Clinical features including duration of hospitalization in NICU, duration of mechanical ventilation and continuous positive airway pressure (CPAP), and total oxygen therapy duration in days were collected. Duration of hospitalization in NICU, mechanical ventilation, continuous positive airway pressure (CPAP), and total oxygen therapy were analysed presence of respiratory distress syndrome (RDS), necrotizing enterocolitis (NEC), bronchopulmonary dysplasia (BPD), sepsis, cardiac disease, resuscitation history, phototherapy, surfactant treatment, extraocular surgery, total parenteral nutrition (TPN), duration of antibiotic use, presence and stage of intraventricular haemorrhage (IVH) were also questioned for each case. Maternal risk factors such as age, presence of gestational diabetes, early membrane rupture, pre-eclampsia, multiple pregnancies, in-vitro fertilization, and antenatal steroid treatment were collected. Even if the received data was not complete, the data sent were included if they were consistent and met the criteria.

Statistical analyses were performed using SPSS v22.0. The normal distribution of the continuous variables was tested using the Kolmogorov–Smirnov test, and the homogeneity of variance was tested using Levene’s test. Descriptive statistics as frequencies, percentages, mean (± standard deviation), and median values (minimum, maximum) were first calculated for all variables of interest. Pearson’s chi-square, Fisher’s exact, likelihood ratio, and McNemar’s tests were used for comparisons of categorical data. Prospective selective multinominal logistic regression analysis was performed to identify important risk factors for the development of aggressive ROP (A-ROP). Statistical significance was set as a 2-tailed p-value < 0.05.

## Results

Data of 376 infants treated between January 2003 and December 2020 from 33 Reference Centres for ROP; 16 university hospitals (137 infants), 15 state hospitals (229 infants), 2 private hospitals (10 infants) were received. Ten patients were excluded due to insufficient data or duplication, and 366 patients were included into the study. The distribution of hospitals where infants were born and received NICU care was given in Table 1. Totally, 78.4% (n=287) of the infants were referred to the reference centres for treatment.

**Table 1.** Demographic data of infants

| Table 1. Demographic data of infants |  |  |
| --- | --- | --- |
|  | Number of patients | Percentage |
| Development status |  |  |
| Developed cities | 266 | 72.7% |
| Underdeveloped cities | 100 | 27.3% |
| Hospital Type |  |  |
| University Hospital | 44 | 12% |
| State Hospital | 75 | 20.5% |
| Private Hospital | 247 | 67.5% |
| Level of NICU |  |  |
| Level 1-2 | 77 | 21% |
| Level 3 | 243 | 66.4% |
| Level 4 | 46 | 12.6% |
| Neonatologist at NICU |  |  |
| Yes | 226 | 66.1% |
| No | 116 | 33.9% |
| ROP screening by |  |  |
| Ophthalmologist in the same hospital | 118 | 32.2% |
| Invited ophthalmologist from another center | 176 | 48.1% |
| Referring the infant to another hospital | 72 | 19.7% |
| Total stay at NICU (n: 363) |  |  |
| 0-2 weeks | 57 | 15.7% |
| 3-4 weeks | 170 | 46.8% |
| 5-8 weeks | 94 | 25.9% |
| >8 weeks | 42 | 11.6% |
| Total oxygen treatment duration (n: 218) |  |  |
| <1 week | 30 | 13.7% |
| 1-3 weeks | 105 | 48.2% |
| 15,>3 weeks | 83 | 38.1% |
| Duration of mechanical ventilation (n: 203) |  |  |
| <1 week | 132 | 65.0% |
| 1-3 weeks | 45 | 22.2% |
| >3 weeks | 26 | 12.8% |
| Duration of CPAP (n: 196) |  |  |
| <1 week | 144 | 73.5% |
| 1-3 weeks | 43 | 21.9% |
| >3 weeks | 9 | 4.6% |

NICU: Neonatal Intensive Care Unit; ROP: Retinopathy of Prematurity; CPAP: Continuous positive airway pressure

The 33 participating centres had ophthalmology units for ROP screening, all of the centres performed laser photocoagulation and/or anti-vascular endothelial growth factor (anti-VEGF) treatments, and one centre performed vitreoretinal surgery.

The mean GA of infants was 33±1 (32 – 37) wk, and the mean BW was 1896 ± 316 g (1505 – 3760 g). The demographic data and the duration and type of oxygen delivery were listed in Table 1. Maternal and prematurity-related comorbidities were presented in Table 2.

**Table 2.**
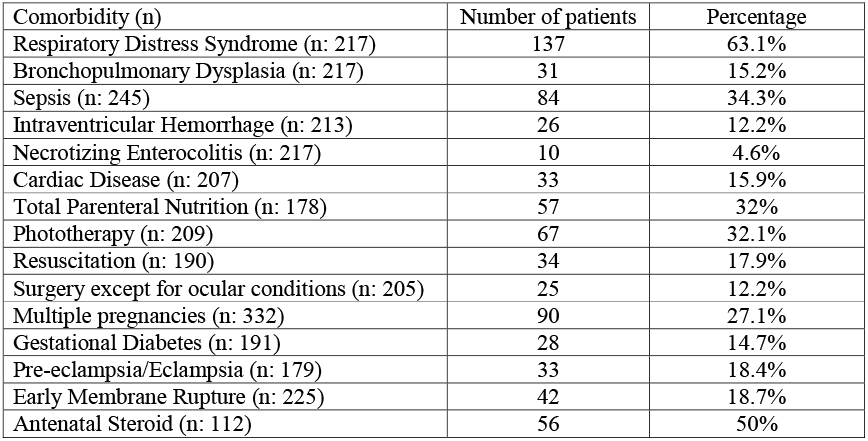
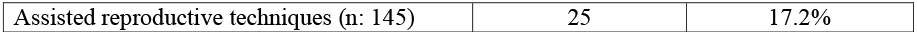
Maternal and prematurity-related comorbidities

| Table 2. Maternal and prematurity-related comorbidities |  |  |
| --- | --- | --- |
| Comorbidity (n) | Number of patients | Percentage |
| Respiratory Distress Syndrome (n: 217) | 137 | 63.1% |
| Bronchopulmonary Dysplasia (n: 217) | 31 | 15.2% |
| Sepsis (n: 245) | 84 | 34.3% |
| Intraventricular Hemorrhage (n: 213) | 26 | 12.2% |
| Necrotizing Enterocolitis (n: 217) | 10 | 4.6% |
| Cardiac Disease (n: 207) | 33 | 15.9% |
| Total Parenteral Nutrition (n: 178) | 57 | 32% |
| Phototherapy (n: 209) | 67 | 32.1% |
| Resuscitation (n: 190) | 34 | 17.9% |
| Surgery except for ocular conditions (n: 205) | 25 | 12.2% |
| Multiple pregnancies (n: 332) | 90 | 27.1% |
| Gestational Diabetes (n: 191) | 28 | 14.7% |
| Pre-eclampsia/Eclampsia (n: 179) | 33 | 18.4% |
| Early Membrane Rupture (n: 225) | 42 | 18.7% |
| Antenatal Steroid (n: 112) | 56 | 50% |
| Assisted reproductive techniques (n: 145) | 25 | 17.2% |

Most of the neonatal care was conducted by private NICUs (67.5%). The rate of private NICUs in developed cities (74.1%) was higher than in underdeveloped cities (50%) (p<0.0001). There was at least one neonatologist in 49% of NICUs regardless of hospital type in the underdeveloped cities and in 73.1% in the developed cities (p<0.0001). However, the rate of ROP examination by an ophthalmologist at the same hospital with NICU was 44% in the underdeveloped cities and 27.8% in the developed cities (p<0.0001). The absence of neonatologists and ophthalmologists was statistically higher in non-university hospitals and lower levels of NICU (p<0.0001 for all). The neonatologist was present in all level 3B and 4 NICUs but in only 68.5% of level 3A NICUs and in 39% of level 1 and 2 NICUs. The rate of presence of a neonatologist in the university hospitals was 95.5% which was statistically significantly higher than that in the state hospitals (84%) and in the private hospitals (54.3%) (p<0.0001). The rate of ROP examination by an ophthalmologist at the same hospital with NICU was 88.6% in the university hospitals, 68% in the state hospitals, and 11.3% in private hospitals (p<0.0001).

In the present study, 294 infants (80.3%) were examined at the appropriate time according to Turkey ROP Screening Guideline which was postnatal 4-5 wk The first examination time was postnatal 6 wk in 32 infants (8.7%) and postnatal 7 wk in 40 infants (10.9%). The first examination timing was statistically significantly later in lower levels of NICU(p=0.001), in the absence of ophthalmologist at the same hospital with NICU (p=0.003), and non-university hospitals (p=0.025). ROP was present in 328 infants (90.9%) at the first examination. The presence of ROP at the first examination was statistically significantly (p<0.05) higher in lower levels of NICU and in the absence of ophthalmologist at the same hospital (Table 3). TR-ROP developed in a mean of 6.16±2.04 wk (4-18 wk). TR-ROP was detected at fourth week in 56 infants (15.4%). ROP type was A-ROP in 60 infants (16.4%) and type 1 ROP in 306 infants (83.6%).

**Table 3.**
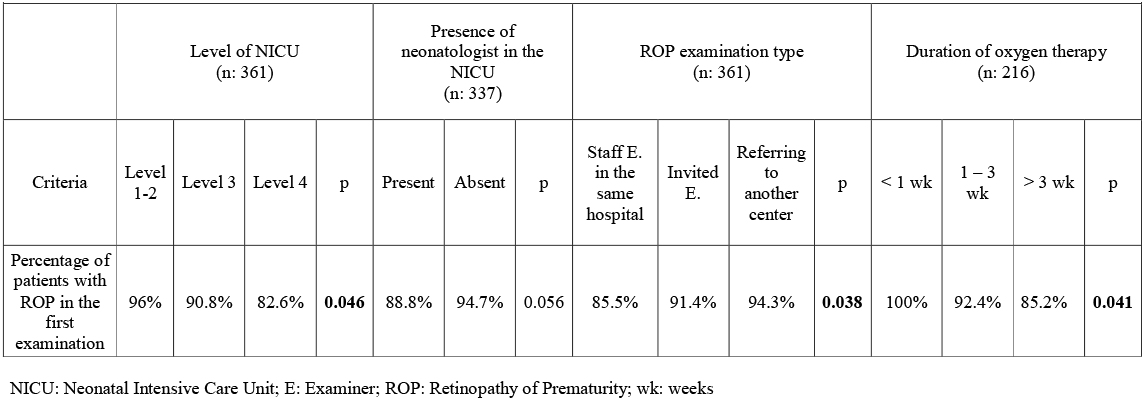
Comparison of NICUs according to the different criteria for the presence of ROP in the first examination.

| Criteria | Level of NICU<br>(n: 361) |  |  |  | Presence of<br>neonatologist in the<br>NICU<br>(n: 337) |  |  | ROP examination type<br>(n: 361) |  |  |  | Duration of oxygen therapy<br>(n: 216) |  |  |  |
| --- | --- | --- | --- | --- | --- | --- | --- | --- | --- | --- | --- | --- | --- | --- | --- |
|  | Level<br>1-2 | Level 3 | Level 4 | p | Present | Absent | p | Staff E.<br>in the<br>same<br>hospital | Invited<br>E. | Referring<br>to<br>another<br>center | p | < 1 wk | 1 – 3<br>wk | > 3 wk | p |
| Percentage of<br>patients with<br>ROP in the<br>first<br>examination | 96% | 90.8% | 82.6% | **0.046** | 88.8% | 94.7% | 0.056 | 85.5% | 91.4% | 94.3% | **0.038** | 100% | 92.4% | 85.2% | **0.041** |

NICU: Neonatal Intensive Care Unit; E: Examiner; ROP: Retinopathy of Prematurity; wk: weeks

The frequency of A-ROP was higher in South-eastern Anatolia than other regions (p<0.001) (Figure 1). The frequency of A-ROP was lower in the state hospitals than university and private hospitals (p=0.037, p=0.001 respectively). The frequency of A-ROP was lower in the

**Figure 1.**
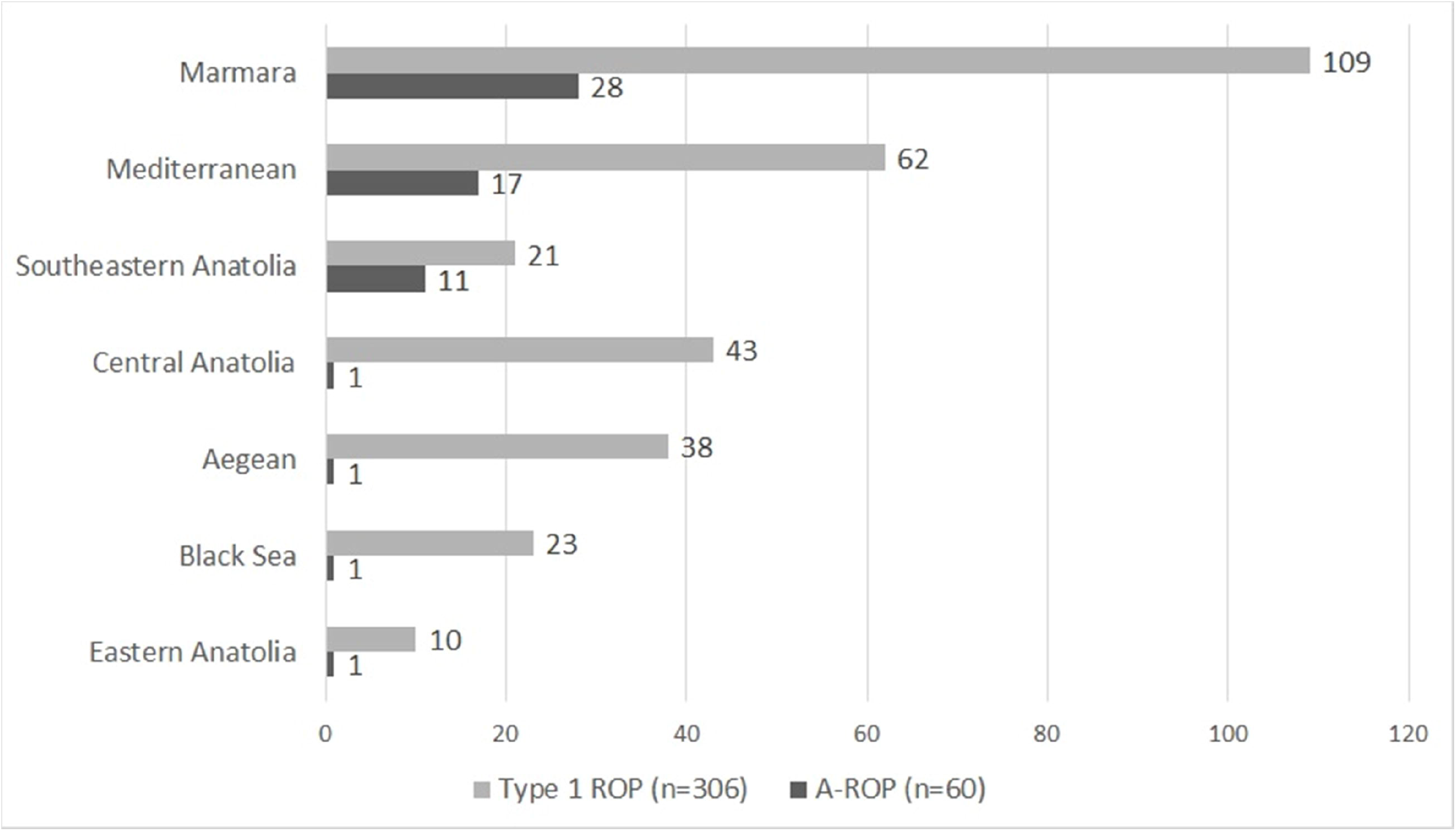
The numbers of aggressive ROP (A-ROP) and type 1 ROP of each region is presented. Although the number of A-ROP cases are higher in Marmara and Mediterranean regions, the rate of A-ROP is statistically higher in Southeast Anatolia Region, where is socio-economically underdeveloped.

Level 4 NICUs than Level 3 NICUs (p=0.016). The comparisons according to the presence of neonatologist and ophthalmologist are presented in Table 4.

**Table 4.** Comparison of (A-ROP) frequency according to the different NICU characteristics

|  |  |  |  |  |  |  |  |  |  |  |  |  |  |  |  |  |  |
| --- | --- | --- | --- | --- | --- | --- | --- | --- | --- | --- | --- | --- | --- | --- | --- | --- | --- |
|  | Development status (n: 366) |  |  | Hospital type (n: 366) |  |  |  | Stage of NICU (n: 366) |  |  |  | The presence of neonatologist (n:342) |  |  | The presence of ophthalmologist (n:366) |  |  |
| Criteria | Developed (n:266) | Under-developed (n:100) | p | University (n:44) | State (n:75) | Private (n:247) | p | Level 1 & 2 (n:77) | Level 3 (n:243) | Level 4 (n:46) | p | Present (n:226) | Absent (n:116) | p | Present (n:118) | Absent (n:248) | p |
| Percentage of A-ROP eyes | 15% | 19% | 0.046 | 16% | 4% | 20% | 0.004 | 16% | 19% | 4% | 0.049 | 17% | 15% | 0.720 | 8% | 21% | 0.003 |

A-ROP: aggressive ROP; NICU: Neonatal intensive care unit

Multinominal logistic regression analysis was performed to identify hospital-related risk factors in the development of A-ROP (Table 5).

**Table 5.** Risk factors for Aggressive ROP development, multinominal logistic regression analysis

| Table 5. Risk factors for Aggressive ROP development, multinomial logistic regression analysis |  |  |  |  |  |  |
| --- | --- | --- | --- | --- | --- | --- |
| | $\beta$ coefficient | Standard error | p | OR | 95.0% CI for OR | |
| Southeastern Anatolia Region | 1.228 | 0.418 | 0.003 | 3.413 | 1.505 | 7.742 |
| Absence of ophthalmologist | 1.209 | 0.387 | 0.002 | 3.350 | 1.568 | 7.154 |
| Constant | -2.689 | 0.363 | <0,001 | 0.068 |  |  |

Dependent variable: A-ROP; Nagelkerke $R^2=0.085$ ; Accurate forecast rate=83.61%
OR: Odds ratio, CI: Confidence interval

## Discussion

It is well known that the incidence and severity of ROP increase with decreasing GA and BW. It is unusual to see a TR-ROP development in infants with a GA of > 28 wk or a BW >1250 g in optimum NICU conditions which is usually available in high-income countries. However, there is a significant number of infants with a GA of >32 wk and BW>1500 g in suboptimal NICU conditions in low to middle-income countries. Our study reveals the demographic and clinical findings of TR-ROP in preterm infants with a GA of >32 wk and BW of >1500 g and aims to point out possible risk factors that need improvement.

Gilbert et al.^1^ reported that more mature infants are developing TR-ROP in low/middle-income countries compared with high-income countries and ROP screening programs should be structured according to the socio-economical characteristics of the countries and their populations. Shah et al.^5^ reported that developing countries such as India are facing a new epidemic similar to the first epidemic of ROP which was related to high oxygen delivery and also called oxygen-induced retinopathy. They named the increasing incidence of TR-ROP in mature and heavier infants as the third epidemic of ROP.^5^ This was confirmed by other authors from China and India.^6,7^ Bas et al.^2^ evaluated the incidence, risk factors, and severity of ROP with a prospective, multicentric study in Turkey and showed that more mature and heavier infants were at risk for TR-ROP. The study included 6115 infants, 1151 of whom (19%) were with a GA>32 wk; any stage of ROP was found in 61 infants (5.3%), and TR-ROP was detected in 6 infants (0.5%). They have suggested that the ROP screening program in Turkey should be expanded to include premature infants with a BW<1700 g or GA<34 wk to detect all infants requiring treatment.^2^

The rate of ROP may be considered as an indicator of the quality of neonatal care.^8^ Bas et al.^2^ reported that the rates of TR-ROP were lower in the university hospital NICUs, where the neonatal care is likely to be better, and reported more mature and heavier infants developing TR-ROP especially in private hospitals. Similarly, in our study, the presence of ROP at the first examination was higher in lower levels of NICU. Shah et al.^5^ indicated that increasing preterm births and survival rates, inadequate oxygen therapy, and failure to comply with the ROP screening program may increase the incidence of ROP in big infants.

Shah et al.^9^ reported loss of retinal vessels from zone II to zone I in relatively mature preterm infants (GA of 31.7 wk [range 28–35 wk]) and progression to A-ROP with 100% oxygen exposure. The number of A-ROP cases was higher in Marmara, Mediterranean Sea, and South-eastern Anatolia regions. The higher number of A-ROP cases in the socioeconomically developed Marmara and Mediterranean Regions may be related to the higher rates of private NICUs in these regions. In our country, in the socioeconomically developed regions, immigration and refugee rates are higher. This situation forces the health system in these regions, may cause more private hospitals to be opened without enough control, and as a result, affects the quality of intensive care in these regions. Further studies are needed to reach more definite conclusion on this topic. For all that, we consider that decreased quality of neonatal care promotes the development of A-ROP in which may be related to oxygen-induced retinopathy. Future studies with detailed information on the follow-up of oxygen saturation level of the neonates are needed to conclude.

Prolonged oxygen therapy was found to be a risk factor for the development of ROP not at the initial examination but at subsequent examinations in the present study. This may be caused by inadequate oxygen delivery or non-compliance to target saturation levels or comorbidities that increase the development of ROP. Lack of neonatologist and shortage of nurses controlling the saturation levels may result in lower quality of NICU service which may lead to the development and progression of ROP and A-ROP which may be linked to a kind of oxygen-induced retinopathy in more mature and heavier preterm infants. In our study, the lack of neonatologists was found to be statistically significantly higher in underdeveloped cities, in lower levels of NICU, and in non-university clinics, especially private hospitals.

Our study included infants without the major risk factors related to GA and BW. Several risk factors such as length of stay in NICU, RDS, BPD, NEC, anaemia, transfusion, sepsis, intracranial haemorrhage, cardiac problems, nutrition, extraocular surgery, and maternal risk factors such as maternal diabetes, pre-eclampsia, assisted conception had been associated with the development of TR-ROP in the literaure.^10-18^ We have evaluated the rates of such risk factors in our patients, and we found similar or lower rates as compared to other cohorts in the literature.^2,19-21^ We did not include patients without TR-ROP, therefore we could not compare the effect of these factors on the development of ROP.

In our study, ROP was present in 90.9% of patients at the first examination which was performed at postnatal fourth week, and TR-ROP developed in a mean period of 6.16 wk This finding may indicate that the onset of ROP corresponds more closely with postmenstrual age than with postnatal age.^22^ ROP onset was reported to begin at roughly 30 wk of PMA and peaked at a PMA of 36–38 wk in infants with BW <1251 g, irrespective of GA at birth.^23^ Our study revealed that ROP was already present at PMA of 36 wk and TR-ROP developed at a mean of 38 wk similarly. The high rates of ROP in the first examination in the present study may suggest that the first ROP examination may be done earlier (even before postnatal 4 wk) in bigger prematures with a BW>1500 g and GA>32 wk and should be followed up closely if there is any ROP.

In our study, it was shown that 72 infants (19.7%) could not be screened for ROP on time. We also found that premature infants had their first examination for ROP later than the ideal time in hospitals where there is no ophthalmologist on charge in the same hospital. In such hospitals, the first and ongoing ROP examinations were performed by an invited ophthalmologist from another centre or with referral of the infant to another hospital for ROP screening. The timing of first ROP examination was postnatal 4-5 wk in 90.7% of the infants in the hospitals where an ophthalmologist was on charge. However, this rate was 75% if there is no ophthalmologist in the same hospital. The presence of ophthalmologist in the same hospital seems to increase the proper conduction of ROP screening by letting timely examinations and preventing delay in detection of ROP. When the ophthalmologist performing the ROP screening is present in the same hospital, both the screening and treatment can be planned readily when needed for even a single infant. However, examinations may tend to be grouped together when it is to be done by an invited ophthalmologist from other hospitals. This may cause some delay in the screening programs e.g. to postpartum 5 to 7 wk instead of 4 wk which may explain the detection of higher rate of ROP at the first examination. Similarly, Mousavi et al. reported that late examination increases the prevalence of ROP and the incidence of serious ROP in preterm infants.^24^ There is also a relationship between absence of ophthalmologist and A-ROP rates which is parallel to the quality of neonatal care. The lack of ophthalmologist was more prominent in non-university clinics and especially in private hospitals in the present study. Our study suggests that hospitals with NICU must have an ophthalmologist trained in ROP screening as well as a neonatologist.

This study has some limitations because of its retrospective nature. First, some of the data about systemic parameters in NICU were lacking in some infants. The most important of these was the lack of oxygen saturation levels as we mentioned above. Second, since only infants with TR-ROP who needed treatment were included in the study, we could not make comparisons for the risk factors for ROP development in these bigger infants. On the other hand, the strength of this study can be listed as follows: This is the first study analysing the regional differences and the effect of presence of the ophthalmologist and neonatologist in the same hospital as NICU on the development of TR-ROP in a nation-wide study in Turkey.

This study emphasizes the high rate of ROP at the first examination in these bigger babies and progression to TR-ROP in a short time. The results of our study suggest the need for a timely (even earlier) screening for bigger infants and a revision to expand the limits of the ROP screening program to bigger infants in at least underdeveloped parts of Turkey. This may be generalized to all of the underdeveloped countries.

In conclusion, more mature and heavier preterm infants may develop TR-ROP especially when the ideal NICU conditions cannot be provided. Screening criteria for ROP need to be revised for the coverage of bigger infants in Turkey depending on the NICU conditions of the hospitals. The ROP in these infants may be more aggressive like A-ROP and may progress rapidly in a short time. The first examination for ROP should be no later than postnatal fourth week with a close follow-up. Increasing the number of well-educated neonatologists and ophthalmologists as well as other NICU conditions will improve neonatal care for ROP to the standards of developed countries.

## Data Availability

All data produced in the present study are available upon reasonable request to the authors

## Data availability statement

Data are available on reasonable request.

## Ethics statements

### Patient consent for publication

Not required.

### Ethics approval

This study was approved by the Ethics Commission of the Gazi University (Approval Number: 2020-191). We adhered to the principles of the Declaration of Helsinki.

## Footnotes

### Contributors

SO and ZOT designed the study. SO and HBO collected the data. HBO labelled the data. SO and HBO verified the raw dataset and labels. SO and HBO developed the algorithm, analysed the data, drafted the manuscript and interpreted the results. All authors contributed to the manuscript revision. All authors approved the submitted version.

### Provenance and peer review

Not commissioned; externally peer reviewed.

